# Association between plasma brain-derived p-tau variants and anxiety phenotypes in older adults

**DOI:** 10.64898/2026.10.02.26364610

**Authors:** Ana Paula Costa, Helmet T. Karim, Marissa F. Farinas, Nya Nicholson, Mary M. Geissinger, Xuemei Zeng, Dana L. Tudorascu, Meryl A. Butters, Thomas K. Karikari, Carmen Andreescu

## Abstract

**Objectives:** To determine whether distinct anxiety phenotypes (rumination, global anxiety and worry; RAW) are associated with plasma brain-derived (BD) phosphorylated-tau (p-tau) biomarkers of Alzheimer’s disease (AD) in older adults.

**Design:** Cross-sectional observational study.

**Methods:** Participants were 89 older adults aged ≥50 years (mean age 64.3±8.8 years) enrolled in the RAW Brain study (R01 MH108509) in Pittsburgh, Pennsylvania. Anxiety phenotypes were assessed using the Penn State Worry Questionnaire (PWSQ≥50), Rumination Subscale from the Response Style Questionnaire (RSQ>50), and Hamilton Anxiety Rating Scale (HARS>17). Participants were classified as High-RAW if they exceeded established severity thresholds on ≥1 domain; continuous anxiety scores were used for primary analyses. Plasma BD-tau, BD-p-tau181, BD-p-tau217, and BD-p-tau231, and matched non-BD versions were quantified using the NULISAseq™ CNS Panel. Associations between anxiety phenotypes and tau biomarkers were examined using multivariable linear regression adjusted for age, sex, and APOE4 status, with Benjamini-Hochberg false discovery rate correction.

**Results:** Greater global anxiety was associated with higher plasma BD-p-tau181 (β=0.308, 95% CI [0.003,0.048], p=0.047). This association was also observed categorically (adjusted Cohen’s d=0.462, p=0.049). Worry and rumination were not associated with BD-p-tau181, and no anxiety phenotypes were associated with BD-tau, BD-p-tau217, BD-p-tau231, or corresponding non-BD alternatives.

**Conclusions:** Greater late-life global anxiety, but not worry or rumination, was selectively associated with higher plasma BD-p-tau181. These findings extend epidemiologic evidence linking anxiety to AD risk by identifying a biologically specific blood-based correlate linking anxiety with AD-related p-tau biology. Longitudinal studies incorporating amyloid and tau biomarker confirmation are needed to determine whether anxiety precedes, accompanies, or results from early AD pathophysiology.

**Highlights:**

**1) What is the primary question addressed by this study? —The question addressed by the study must limited to only one sentence.** Is late-life anxiety, including rumination, global anxiety, and worry, associated with plasma brain-derived (BD) phosphorylated-tau (p-tau) biomarkers of Alzheimer’s disease in older adults?
**2) What is the main finding of this study? —The finding must be limited to two sentences.** Greater global anxiety, but not worry or rumination, was associated with higher plasma BD-p-tau181 after adjustment for age, sex, and APOE4 status. No associations were observed with other BD-tau variants or with non-BD-tau versions.
**3) What is the meaning of the finding? —The meaning of the finding must be limited to one sentence.** These findings suggest that global anxiety may be linked to early Alzheimer’s disease-related tau biology and support further longitudinal investigation of anxiety as a potential marker of preclinical disease.

## INTRODUCTION

Late-life anxiety is one of the most common psychiatric conditions affecting older adults and is increasingly recognized as being associated with cognitive decline, Alzheimer’s disease (AD), and related dementias^1–3^. Beyond its well-established impact on quality of life, functional impairment, and increased healthcare utilization^4^, prospective cohort studies and meta-analyses have consistently shown that anxiety is associated with 77% higher risk of cognitive impairment^1^, 53% higher risk of Alzheimer’s disease^5^, and 65% higher risk of vascular dementia^6^. However, because much of the available evidence derives from populations that already exhibit cognitive symptoms or biomarker evidence of neurodegeneration^7^, it remains unclear whether anxiety constitutes a risk factor for decline^4,8^, an early manifestation of it^1,9,10^, or both. Clarifying this relationship has become increasingly important as blood-based biomarkers now permit the investigation of AD-related biological processes during the preclinical stages of disease.

Recent studies on dementia prevention estimate that approximately 45% of dementia cases are linked to a combination of potential risk factors, including cardiovascular risk factors, high cholesterol, depression, and lower education attainment^11^. Anxiety has also been recognized as one of these potentially modifiable dementia risk factors^6,11,12^. However, despite growing recognition of its clinical importance, anxiety is typically treated as an umbrella term used to cover various categorical disorders as well as heterogenous clusters of symptoms assessed by various questionnaires^13^. This broad conceptualization overlooks the biological heterogeneity of anxiety, which comprises multiple dimensions with distinct cognitive, behavioral, and neurobiological characteristics^14,15^. Rumination, global anxiety, and worry (RAW) are three distinct and highly prevalent anxiety phenotypes. Our group has shown that these dimensions exhibit dissociable behavioral and neurobiological profiles^15–18^, suggesting that combining these constructs could mask meaningful phenotype-specific associations with AD-related processes.

Despite accumulating epidemiological evidence linking late-life anxiety to an increased risk of dementia, the biological mechanisms underlying this association remain poorly understood. Neuroimaging and biomarker studies have reported associations between anxiety and several markers of AD pathology. Higher amyloid-β (Aβ) burden predicts increasing anxiety symptoms over time in cognitively normal older adults^19^. Subcortical amyloid deposition correlates with self-reported anxiety, an association that is stronger in APOE4 carriers^20^. In amnestic mild cognitive impairment (MCI), anxiety severity is associated with faster medial temporal atrophy and predicts conversion to AD^21^. A meta-analysis of prospective cohort studies found that anxiety predicts incident cognitive impairment and dementia in community samples, although it did not predict conversion to dementia among memory-clinic patients already diagnosed with MCI^1^. Other investigations report weak or no relationships, particularly for Aβ and tau^22,23^. These inconsistencies likely reflect substantial heterogeneity across studies, including differences in participant characteristics, disease stage, anxiety phenotyping, and the biomarkers used to quantify AD pathology. In particular, most studies have relied on conventional plasma tau assays that do not distinguish tau originating from the central nervous system (CNS) from the peripherally expressed “big tau” isoform, thereby limiting their specificity for brain tau pathology^24,25^.

Recent advances in ultrasensitive blood-based biomarkers have substantially improved the ability to detect AD-related pathology during the preclinical stage. Among these developments, brain-derived tau (BD-tau) assays represent an important advance because they selectively quantify tau originating from the central nervous system while excluding the peripherally expressed “big tau” isoform that dominates conventional plasma measurements. BD-tau and its phosphorylated variants are strongly associated with levels of cerebrospinal fluid (CSF) biomarkers and neuropathological evidence of AD and may therefore provide a more specific indicator of neuronal tau pathology than traditional plasma tau measurements^25^. Whether anxiety phenotypes are differentially associated with these brain-derived tau biomarkers has not previously been investigated.

The present study examined the relationships between dimensional measures of rumination, global anxiety, and worry and plasma BD-tau variants in older adults enrolled in the RAW Brain study, a cohort enriched for anxiety-related symptoms. We focused on anxiety dimensions rather than categorical psychiatric diagnoses to capture the full spectrum of symptom severity and determine whether specific anxiety phenotypes demonstrate distinct associations with early AD-related biological processes. We hypothesized that greater worry and global anxiety would be associated with higher concentrations of brain-derived phosphorylated tau (BD-p-tau), after adjustment for age, sex, and APOE4 carrier status and that these relationships would be specific to brain-derived rather than conventional plasma tau variants.

## METHODS AND MATERIALS

### Participants and study design

The study cohort consisted of participants from the RAW Brain study (R01 MH108509 The RAW Brain – The Effect of Rumination, Anxiety and Worry on Aging and Dementia Risk) in Pittsburgh, Pennsylvania, USA. The RAW Brain study is an ongoing observational longitudinal study with visits 2 years apart including demographic, clinical, behavioral, plasma biomarkers, and neuroimaging data collection; aimed at understanding the relationships between late-life anxiety and Alzheimer’s disease and related dementias (ADRD) risk. Participants >50 years of age were recruited through a web-based recruitment resource at the University of Pittsburgh (Pitt+Me), in-person referrals, flyers, and radio/television ads. For this study, we included the baseline assessment of 89 participants. Participants were recruited using key dimensions of anxiety - rumination, global anxiety, worry (RAW) - this dimensional design allowed us to tap into an understudied yet highly prevalent population of older participants with severe rumination, anxiety or worry, that may or may not qualify for a categorical diagnosis^26,27^. Participants could have a diagnosis of anxiety disorder (e.g., generalized anxiety disorder, panic disorder, etc.) and/or depressive disorders (e.g., major depressive disorder, persistent depressive disorder, etc.), but that was not required. Exclusion criteria for the study were 1) other major psychiatric disorder including any form of psychosis, bipolar disorder, autism spectrum disorder, and intellectual development disorder; 2) dementia [Modified Mini-Mental State Examination score <84]; 3) high suicide risk; 4) use of selective serotonin reuptake inhibitor or serotonin and norepinephrine reuptake inhibitor antidepressants that participants were not willing/able to discontinue before the scan (5- to 14-day washout depending on the antidepressant, 6 weeks for fluoxetine); 5) history of drug or alcohol abuse within the last 6 months; 6) high doses of benzodiazepines (≥2 mg of lorazepam/day); 7) uncorrected vision problems that preclude testing; 8) below sixth grade reading level; 9) clinical diagnosis of cerebrovascular accident, multiple sclerosis, vasculitis, or significant head trauma; and 10) contraindications to MRI. The University of Pittsburgh Institutional Review board approved the study. All participants provided written informed consent.

### Anxiety phenotype classification and measures

Along with demographic information (age, sex, race, ethnicity, and education), we assessed the following: worry severity (PSWQ, Penn State Worry Questionnaire^28,29^), global anxiety (HARS, Hamilton Anxiety Rating Scale^30^), and rumination (RSQ, Rumination Subscale from the Response Style Questionnaire^31^). Participants were classified as High-RAW (n=36) if they meet thresholds on ≥1 domain: PSWQ ≥50 and/or RSQ >50, and/or HARS >17. Participants who did not meet any of the above thresholds were classified as Low-RAW (n=53). For the primary regression analyses, all three measures were treated as continuous variables to preserve statistical power and capture dimensional variation in anxiety phenotypes.

Participants were further characterized clinically using measures of depression (HAMD, Hamilton Depression Rating Scale), neuroticism (FFI, Five-Factor Inventory), perceived stress (Perceived Stress Scale – 10 item), medical burden (CIRS-G, Cumulative Illness Rating Scale – Geriatrics), functional impairment (WHODAS 2.0, World Health Organization Disability Assessment Schedule II-36 Items), lifetime history of psychiatric disorders as per DMS-5 criteria (MINI, Mini International Neuropsychiatric Interview) as well as a comprehensive neuropsychological assessment. Global cognitive function was assessed with the Repeatable Battery for the Assessment of Neuropsychological Status (RBANS) and the Montreal Cognitive Assessment (MoCA). The RBANS is a standardized, age-normed test that assesses five domains including immediate and delayed memory, visuospatial ability, language function, and attention and yields a Total Index Score (RBANS total)^32^. The MoCA is a 30-point cognitive screening instrument designed to detect dementia for clinical care and research, it consists of 12 brief tasks covering visuospatial/executive function, naming, memory, attention, language, delayed recall, and orientation that yield a single total score^33^.

### Blood collection and processing

Blood samples were collected according to the procedures described in Zeng et al^34^. Briefly, blood was collected via venipuncture by experienced nurses trained at the Western Psychiatric Hospital – UPMC Clinic. Whole blood from each participant was collected into a 10 mL lavender top EDTA tube. Following each blood draw, the EDTA tubes were promptly inverted 8-10 times and centrifuged at 2,000 *x g* for 10 minutes at 4°C. The resulting plasma samples were aliquoted into cryovials and frozen at −80°C until further analysis.

### NULISAseq assay procedures and data processing

Plasma samples were thawed and centrifuged at 10,000 *x g* for 10 minutes to remove particulates. The supernatants were then analyzed using the NULISAseq™ CNS Disease Panel 120 on an Alamar ARGO™ system as previously described^35^. Briefly, samples were incubated with a cocktail of paired capture and detection antibodies for the included target protein biomarkers and the internal control (IC). The capture antibodies were conjugated with partially double-stranded DNA containing a poly-A tail and a target-specific barcode, while detection antibodies were conjugated with another partially double-stranded DNA containing a biotin group and a matching target-specific barcode. After incubation, the mixtures underwent magnetic bead-based capture, wash, release, recapture, and a second round of wash processes to purify the formed immunocomplexes. A ligation mix, including T4 DNA ligase and a specific DNA ligator sequence, was utilized to ligate the proximal ends of DNA attached to the paired antibodies, generating DNA reporter molecules containing unique target and sample-specific barcodes. The reporter DNA levels were then quantified by Next-Generation Sequencing (NGS). The plasma samples were randomized in two plates for the assay. Three assay controls were run side-by-side with samples for each plate, including the sample control (2 replicates/plate), the inter-plate control (IPC; 3 replicates/plate), and the negative control (2–3replicates/plate). In the case of each BD vs. total p-tau analyte, an identical capture antibody specific for the phospho-epitopes at threonine-181, 217, or 231 was paired with a detection antibody selective for CNS-predominant tau isoforms vs. one that binds all tau isoforms, thus enabling the measurement of brain-derived (BD)- and total-tau species.

Data normalization was performed to remove potential unwanted technical variation. First, IC-based normalization was done by dividing the target counts for each sample well by that well’s IC counts. IPC normalization was achieved by dividing IC-normalized counts by target-specific medians of the IPC (pooled plasma) sample replicates on that plate. Finally, the data was rescaled and log2-transformed to give a more normal distribution for subsequent statistical analyses. These values are hereafter referred to as NULISA Protein Quantification (NPQ) units. The fold change difference between two groups were calculated as 2^(difference^ ^in^ ^NPQ)^. The plate-specific limit of detection (LOD) was calculated for each target assay by taking the mean plus three times the standard deviation (SD) of the unlogged normalized counts for the negative control samples on the plate. LODs were then rescaled and log2-transformed as above. Measurements for sample controls were used to evaluate the reproducibility of the assays, including both the within-run and between-run coefficients of variation (CVs).

### Statistical analysis

Continuous variables were summarized as mean ± SD and compared between Low-RAW and High-RAW groups using the Wilcoxon rank-sum test. Categorical variables were summarized as n (%) and compared using Pearson’s chi-squared test (**Table 1**). Because raw NPQ values for several tau markers departed from normality (Shapiro-Wilk p<0.05), BD-tau variants and non-BD-tau alternatives were transformed using the *bestNormalize* package in R, which selects a normalizing transformation for each analyte through repeated cross-validation and rescales the output to a standard normal distribution. Group differences in tau biomarkers between Low-RAW and High-RAW participants were tested with one-way ANOVA (unadjusted) and ANCOVA adjusted for age, sex, and APOE4 carrier status. Effect sizes are reported as Cohen’s d with 95% CI, and post hoc power was estimated from the observed d assuming a two-sample t-test with unequal group sizes. Convergent validity between each BD-tau variants and its matched plasma versions was assessed with Pearson correlations on the standardized scale. The primary analyses were multivariable linear regression models, one per tau marker, with PSWQ (worry), RSQ (rumination), and HARS (anxiety) entered simultaneously as continuous predictors and adjusted for age, sex, and APOE4 carrier status. Standardized coefficients (β) were obtained by refitting each model on z-scored predictors and outcome. Multicollinearity among model predictors was assessed using variance inflation factors (VIFs), with VIF values <10 considered acceptable. Within each model, p-values were adjusted for multiple comparisons using the Benjamini–Hochberg false discovery rate (BH-FDR) procedure. As a complementary analysis, each anxiety phenotype was also dichotomized at its established clinical cutoff (PSWQ≥50 and/or RSQ>50, and/or HARS>17). Adjusted Cohen’s *d* for each phenotype-marker association was derived from the corresponding model *t*-statistic after adjustment for age, sex, and APOE4 carrier status, with BH-FDR correction applied within each model. APOE4-stratified analyses were not performed because of the limited sample size within APOE4 carrier groups, which would have resulted in insufficient group sizes for reliable, stratified estimates. As a sensitivity analysis, both the RAW comparison and continuous anxiety-phenotype regressions were repeated among participants with data available to derive duration of symptom duration (n=43). Participants were stratified according to symptom duration as <10 years versus ≥10 years, based on the maximum duration code recorded across the PSWQ, RSQ, and HARS duration items. All analyses were performed in R, using the *dplyr, gtsummary, broom, bestNormalize, effectsize*, and *pwr* packages.

**Table 1.** Demographics and Clinical Characteristics of Participants.

| **Table 1. Demographics and Clinical Characteristics of Participants** |  |  |  |  |
| --- | --- | --- | --- | --- |
| **Variable** | **Low-RAW**<br>**N = 53<sup>1</sup>** | **High-RAW**<br>**N = 36<sup>1</sup>** | **p-value<sup>2</sup>** | **Overall**<br>**N = 89<sup>1</sup>** |
| **Age, years, mean (SD)** | 66.6 ± 8.5 | 60.9 ± 8.1 | **0.002** | 64.3 ± 8.8 |
| **Sex, n, %** |  |  | 0.255 |  |
| Male | 19 (36%) | 8 (22%) |  | 27 (30%) |
| Female | 34 (64%) | 28 (78%) |  | 62 (70%) |
| **Race, n, %** |  |  | >0.999 |  |
| White | 45 (85%) | 30 (83%) |  | 75 (84%) |
| Non-White | 8 (15%) | 6 (17%) |  | 14 (16%) |
| **Ethnicity, n, %** |  |  | 0.845 |  |
| Not Hispanic or Latino | 53 (100%) | 35 (97%) |  | 88 (99%) |
| Hispanic or Latino | 0 (0%) | 1 (2.8%) |  | 1 (1.1%) |
| **Education, years, mean (SD)** | 16.8 ± 2.2 | 16.3 ± 2.1 | 0.330 | 16.6 ± 2.2 |
| **APOE4 carrier status, n, %** |  |  | 0.532 |  |
| Non-carrier | 40 (75%) | 30 (83%) |  | 70 (79%) |
| Carrier | 13 (25%) | 6 (17%) |  | 19 (21%) |
| **PSWQ (Worry), mean (SD)** | 31.5 ± 9.3 | 55.1 ± 10.7 | **<0.001** | 41.1 ± 15.2 |
| **RSQ (Rumination), mean (SD)** | 29.3 ± 5.7 | 47.4 ± 12.1 | **<0.001** | 36.7 ± 12.5 |
| **HARS (Anxiety), mean (SD)** | 5.4 ± 3.6 | 15.1 ± 7.0 | **<0.001** | 9.3 ± 7.1 |
| **RBANS total, mean (SD)** | 129.6 ± 133.9 | 107.8 ± 15.0 | 0.514 | 121.1 ± 105.2 |
| **MoCA total, mean (SD)** | 27.3 ± 1.9 | 27.1 ± 2.3 | >0.999 | 27.2 ± 2.1 |
| **BD-total-tau, NPQ, mean (SD)** | 12.8 ± 0.3 | 12.7 ± 0.2 | 0.625 | 12.7 ± 0.2 |
| **BD-p-tau181, NPQ, mean (SD)** | 13.4 ± 0.6 | 13.3 ± 0.5 | 0.760 | 13.4 ± 0.6 |
| **BD-p-tau217, NPQ, mean (SD)** | 11.8 ± 0.4 | 11.7 ± 0.4 | 0.710 | 11.7 ± 0.4 |
| **BD-p-tau231, NPQ, mean (SD)** | 13.1 ± 0.3 | 13.1 ± 0.2 | 0.475 | 13.1 ± 0.2 |

<sup>1</sup> Mean ± SD; n (%)
<sup>2</sup> Wilcoxon rank sum test; Pearson's Chi-squared test
Mean ± SD for continuous variables. n (%) for categorical variables.
p-values: Wilcoxon rank-sum test (continuous). Pearson Chi-squared test (categorical).

## RESULTS

### Participant characteristics

The demographics and clinical characteristics of the participants are described in **Table 1**. A total of 89 participants from the RAW Brain study were included in this cross-sectional analysis. The mean age was 64.3±8.8 years (70% female and 84% white). The High-RAW group (n=36) was significantly younger than the Low-RAW group (n=53; 60.9±8.1 vs. 66.6±8.5 years; p=0.002). Groups did not differ significantly in sex, race, ethnicity, years of education, or APOE4 carrier status. By design, the High-RAW group evidenced markedly elevated scores on all three anxiety phenotype measures compared to the Low-RAW group: PSWQ (55.1±10.7 vs. 31.5±9.3; worry), RSQ (47.4±12.1 vs. 29.3±5.7; rumination), and HARS (15.1±7.0 vs. 5.4±3.6; anxiety; all p<0.001). For plasma biomarkers, brain-derived (BD) tau forms (BD-tau, BD-p-tau181, BD-p-tau217, and BD-p-231) were not significantly different between High- and Low-RAW groups. The RBANS total score and MoCA total score were not significantly different between High- and Low-RAW groups.

### Plasma brain-derived tau variants by anxiety phenotypes

Participants were stratified by High- and Low-RAW groups. None of the plasma BD-tau or BD-p-tau forms differed significantly between the two groups, either in unadjusted comparisons or after adjusting for age, sex, and APOE4 carrier status (**Figure 1**). The non-brain-derived (non-BD) plasma tau or p-tau versions followed the same pattern, with no significant group difference under either model (**Supplementary Figure S1**).

**Figure 1.**
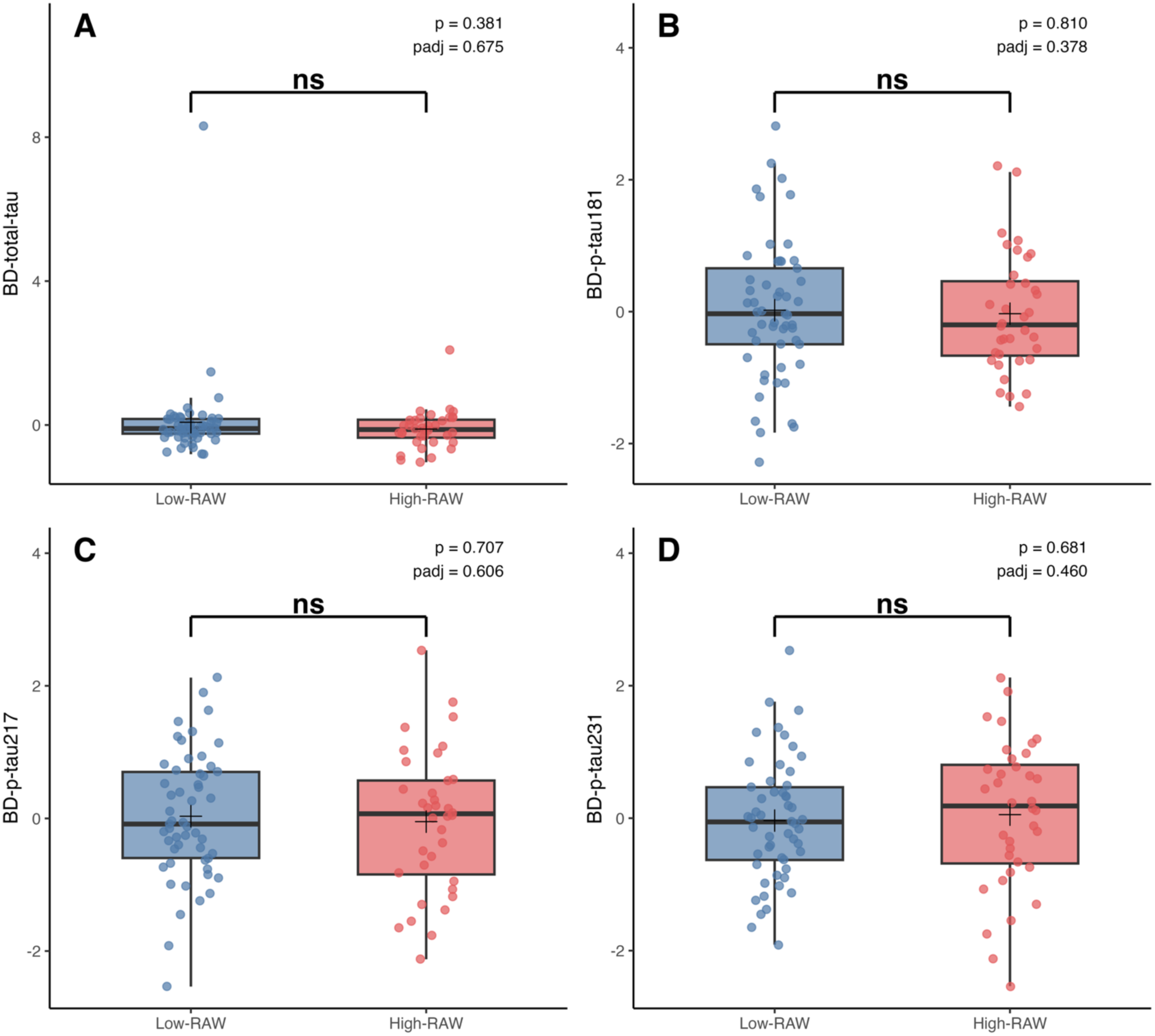
Brain-derived (BD) tau forms between Low- and High-RAW groups. Box plots compare normalized plasma levels of four brain-derived (BD) tau forms between participants stratified into Low-RAW and High-RAW groups (n=89): **(A)** BD-total-tau, **(B)** BD-p-tau181, **(C)** BD-p-tau217, and **(D)** BD-p-tau231; unadjusted (p) and adjusting (padj) for age, sex, and APOE4 carrier status.

Each BD-tau form correlated positively and significantly with its matched non-BD-tau alternative (**Supplementary Figure S2A**). The correspondence was strongest for p-tau217 (rho= 0.79), and p-tau181 (rho= 0.77), and more moderate for p-tau231 (rho= 0.57) and total-tau (rho= 0.41); all p<0.001 (**Supplementary Figure S2B**).

### Associations between anxiety phenotypes and BD-phospho-tau forms

In adjusted linear models (age, sex, and APOE4 carrier status; n=89), greater anxiety was associated with higher levels of BD-p-tau181 (β=0.308, 95% CI [0.003,0.048], p=0.047; **Figure 2**). Worry and rumination showed no association with BD-p-tau181 (both p>0.170), and none of the three anxiety phenotypes were associated with BD-tau, BD-p-tau217, or BD-p-tau231 (**Figure S3A-C**). The same pattern held in the Cohen’s analysis: anxiety in High-RAW vs. Low-RAW was significant for BD-p-tau181 (*d*=0.462, 95% CI [0.03, 0.89], p=0.049; **Figure 3**). None of the non-BD-tau alternatives reached significance for any anxiety phenotypes (**Supplementary Figure S4**).

**Figure 2.**
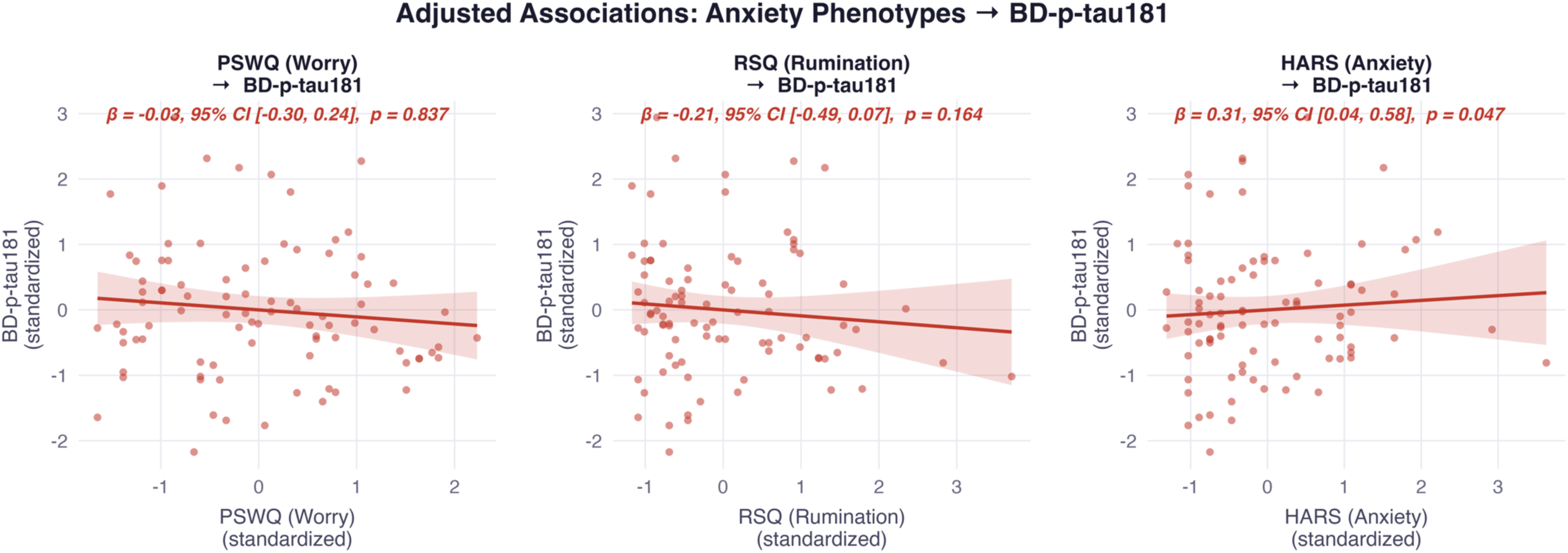
Anxiety-associated phenotypes and their brain-derived tau correlates. The panel corresponds to brain-derived (BD) phosphorylated tau (p-tau) 181 associated with anxiety phenotypes (worry, rumination, and global anxiety). Scatterplots of the BD-p-tau181 against anxiety phenotypes, with linear regression fit (solid line) and 95% confidence band (shaded). All models were adjusted for age, sex, and APOE4 status (N=89); standardized β, 95% CI, and FDR-adjusted p-value are shown.

**Figure 3.**
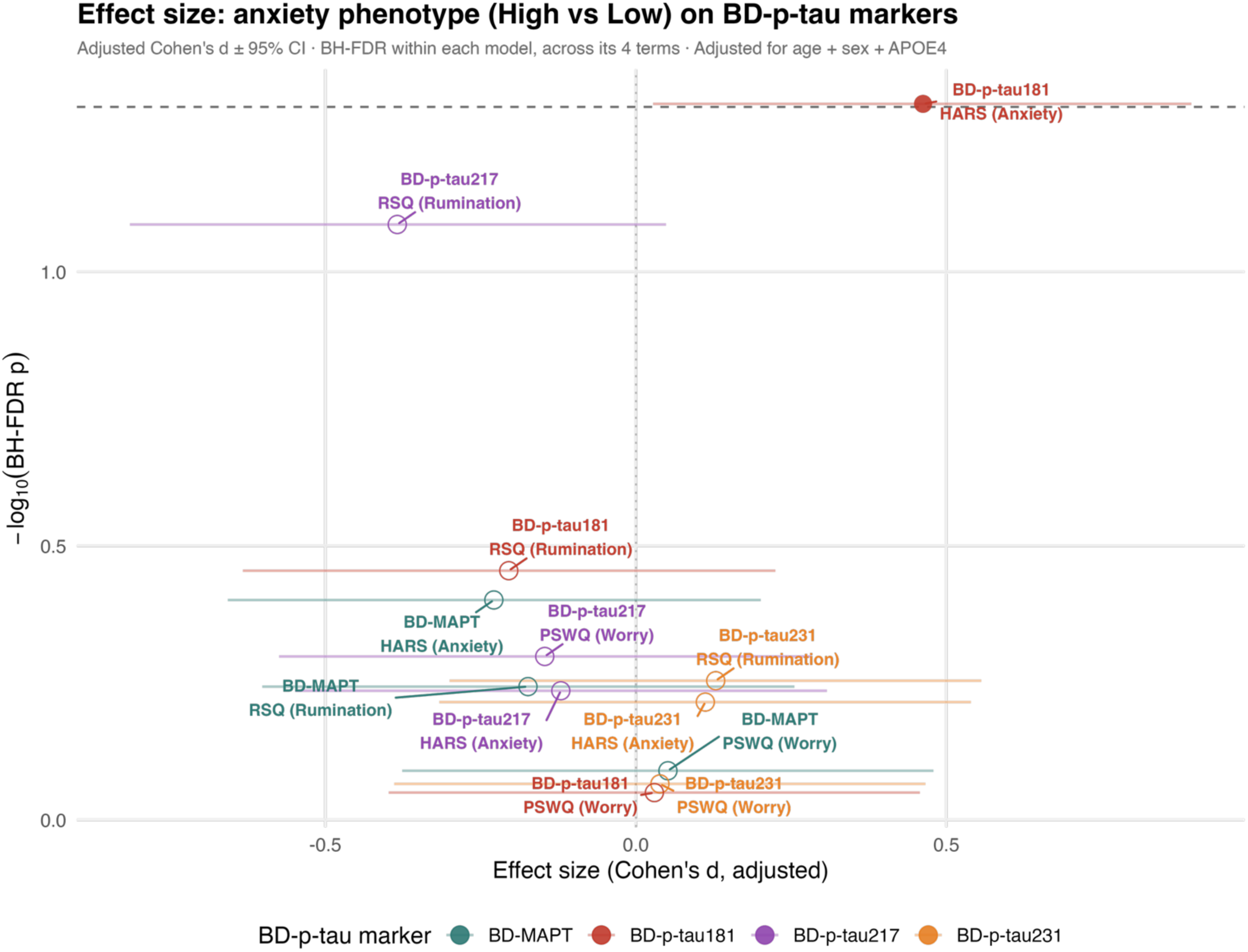
Effect size of High- vs. Low-RAW on BD-p-tau forms. Adjusted Cohen’s d (points) and 95% CI (horizontal lines) for the high-versus-low comparison on each anxiety phenotype by BD-p-tau form, controlling for age, sex, and APOE4 status (n=89 for each comparison; group sizes vary by phenotype threshold). The y-axis shows −log₁₀ of the Benjamini-Hochberg FDR-corrected p value, computed within each phenotype-by-marker model across its four terms (age, sex, APOE4 carries status, group); the dashed horizontal line marks p=0.05.

### Sensitivity and Supplementary Analyses

Sex-stratified analyses were conducted among all 89 participants (**Table 1**). Within each sex, associations between continuous anxiety phenotypes and BD-tau forms (**Supplementary Figure S5A**) and non-BD-tau alternatives (**Supplementary Figure S5B**) did not remain significant after FDR correction. The association between anxiety (HARS) and BD-p-tau181 observed in the full sample was stronger in women (β=0.336, 95% CI [-0.01, 0.68], p=0.141) than in men (β=0.073, 95% CI [-.045, 0.59], p=0.773), suggesting that the full-sample association may be primarily driven by women. However, neither sex-stratified association was statistically significant after FRD correction. Among men, rumination (RSQ) was associated with BD-p-tau231 before FRD correction (β=0.516, 95% CI [0.04, 1.00], p=0.036) but did not survive within-model FDR correction (p=0.082).

Symptom-duration data were available for 43 of the 89 participants (36 High-RAW, 7 Low-RAW), of whom 16 reported symptom duration <10 years and 27 reported ≥10 years (**Supplementary Table S1**). In analyses stratified by symptom duration, RAW group differences in BD-tau variants were not significant after FDR correction (all p≥0.253). Similarly, associations between continuous anxiety-phenotype and BD-tau (**Supplementary Figure S5A**) or non-BD-tau versions (**Supplementary Figure S5B**, all p≥0.24). The HARS-BD-p-tau181 association remained positive in both duration groups (β=0.426, 95% CI [-0.29, 1.115], for <10 years, and β=0.325, 95% CI [-0.24, 0.89], for ≥10 years), although neither association reach significance. Given the small sample size within symptom duration strata, particularly after stratification, these results should be considered exploratory.

## DISCUSSION

In this cross-sectional study of older adults enriched for anxiety-related symptoms, including rumination, global anxiety, and worry (RAW), we examined whether specific anxiety phenotypes related to brain-derived (BD) phosphorylated tau (p-tau) forms measured in plasma. Greater global anxiety, measured by HARS, was associated with higher BD-p-tau181, an association that held after adjusting for age, sex, and APOE4 carrier status and emerged consistently whether anxiety was modeled as a continuous variable (severity of symptoms) or as a dichotomized High-vs. Low-RAW group. Worry and rumination showed no association with any BD-tau variants, and none of the three anxiety phenotypes were related to the other three BD-tau forms (BD-tau, BD-p-tau217, BD-p-tau231) or to any non-BD-tau alternatives (total-tau, p-tau181, p-tau217, p-tau231). This selective pattern, in which global anxiety, rather than worry and rumination, was linked specifically to BD-p-tau181, one of the first plasma phospho-tau epitopes validated as an early Alzheimer’s disease (AD) biomarker^36–39^ and one also liked to AD pathology in cerebrospinal fluid (CSF)^40^, raises the possibility that anxiety severity may track p-tau-related pathophysiological processes earlier in the disease progression than previously shown. This interpretation cannot be determined in a cross-sectional, non-diagnostic sample and will require testing in longitudinal cohorts with amyloid and tau biomarker staging.

A key analytic challenge for blood-based biomarkers of neurological diseases is that most of the circulating proteins are not exclusively brain-derived^41,42^, as peripheral tissue expression and release can contribute to circulating concentrations, confounding assays intended to capture brain-specific pathology. Gonzalez-Ortiz et al. (2023) developed a blood-based assay referred to as BD-tau, using a capture antibody selective for an epitope enriched in the BD-tau isoform, thereby excluding the peripherally expressed “big tau” isoform that dominates unselected plasma total-tau measurements^25^, an approach that also has proven clinical utility to other neurological disorders^43–46^.

More recently, the BD-tau framework has successfully been applied to plasma p-tau resulting in central nervous system (CNS)-selective versions of these biomarkers. Our results, using measures of BD-tau and BD-p-tau and matched non-BD versions from NULISAseq™ proteomics, are consistent with plasma BD-p-tau reflecting CNS-derived signal rather than a purely peripheral one. Each BD-p-tau form correlated positively with its matched non-BD alternatives, with the strongest correlations for p-tau217 and p-tau181 and more moderate for total-tau and p-tau231. This pattern is consistent with prior reports of different coupling between brain and peripheral tau across phosphorylated species, although correlation between matched variants alone does not establish CNS specificity on its own. We speculate that more direct evidence for specificity comes from our finding, where the anxiety association was restricted specifically to the BD-p-tau version of p-tau181, with the non-BD-tau version showing no association with any anxiety phenotypes. The specificity of our finding to the CNS-selective biomarkers supports a CNS origin rather than a generalized peripheral process.

Our results were also specific across anxiety phenotypes. Only global anxiety was associated with plasma BD-p-tau181, worry and rumination were not, despite established literature linking worry to cognitive decline and AD-related pathophysiology in older adults^12,47,48^. These results extend a pattern our group has previously reported, in which worry, rumination, and global anxiety carry distinct rather than interchangeable neurobiological correlates^49^. A plausible physiological account for why global anxiety, specifically, tracked BD-p-tau181 may involve the hypothalamic-pituitary-adrenal (HPA) axis. Late-life generalized anxiety disorder (GAD) is itself characterized by elevated basal and peak salivary cortisol relative to non-anxious older adults^50^. Chronic HPA-axis activation has been proposed as a mechanistic pathway linking psychological stress to Alzheimer disease pathology^51^. Glucocorticoid administration increases both amyloid-β (Aβ) and tau pathology in a transgenic AD mouse model^52^, and chronic stress exacerbates tau hyperphosphorylation and tau-related neurodegeneration through a corticotropin-releasing factor (CRF) receptor-dependent pathway, distinct from and additive to direct glucocorticoid signaling^53^. HARS weights autonomic and somatic symptoms of anxiety, such as palpitations, tension, and sleep disturbance, more heavily than PSWQ or RSQ. Our findings support the HPA-axis dysregulation as the physiological substrate linking anxiety to tau pathology, a measure more closely coupled to autonomic arousal would be expected to track tau phosphorylation more closely than measures of worry or rumination. This is consistent with earlier CSF-based work in a mixed mild cognitive impairment (MCI) sample, in which informant-rated anxiety, and not depression or apathy, was the only neuropsychiatric symptom associated with elevated CSF total-tau^54^. Our result extends that observation to a blood-based, brain-derived tau marker in a non-demented, dimensionally anxious sample. We present the HPA-axis account as a hypothesis consistent with the specificity of our findings, not as a mechanism this cross-sectional study was designed to test; we did not measure cortisol or other HPA-axis markers, and confirming this pathway in humans will require studies that do.

Exploratory sensitivity analyses stratified by sex and symptom duration are reported in full in the Supplementary data. Within each sex, anxiety-phenotype associations with BD-tau and non-BD-tau markers remained non-significant after FDR correction. The HARS–BD-p-tau181 association observed in the full sample was stronger in women than in men, suggesting that the overall association may be primarily driven by women, although it did not reach statistical significance in either sex after FDR correction. Among men, rumination was nominally associated with BD-p-tau23. In the subset with available symptom-duration data (n= 43), neither the RAW group differences nor anxiety-phenotype associations with tau markers were significant after FDR correction. However, the HARS–BD-p-tau181 association remained positive in both symptom-duration strata, consistent with the direction observed in the full sample. Given the small subgroup sample sizes, these stratified analyses should be considered exploratory.

This study has several limitations. First, the design is cross-sectional and cannot establish whether anxiety precedes tau-related change, whether early tau pathology manifests partly as anxiety symptoms, or whether both share an upstream cause. This directionality question is explicitly unresolved even in the largest anxiety-dementia meta-analyses^2,3,6^. Second, we tested numerous phenotype-marker combinations across the primary and sensitivity analyses. Although we corrected for multiple comparisons within each model, only one association survived, and it needs independent replication before any firm conclusion can be drawn. Third, all measures in this study, including BD-p-tau181, anxiety phenotypes, and covariates, were collected at a single baseline visit. We did not have concurrent amyloid or tau PET imaging, CSF biomarkers, or cognitive follow-up data. As a result, we cannot determine the pathological stage represented by elevated BD-p-tau181, nor whether this biomarker predicts future cognitive decline. This is, to our knowledge, the first study to examine the association between BD-tau forms and anxiety phenotypes. A logical step is a longitudinal analysis using the RAW Brain study 2-year follow-up that tracks these markers over time alongside cognitive and imaging outcomes, which would directly address the questions our cross-sectional design cannot answer though this data is not currently available.

Despite these limitations, this study identifies a biologically specific association between late-life global anxiety and a brain-derived plasma biomarker of AD pathology. These findings extend epidemiologic evidence linking anxiety to dementia risk by demonstrating an association with BD-p-tau181 in older adults and suggest that anxiety dimensions may differ in their relevance to AD-related biological processes. Longitudinal follow-up of the RAW Brain cohort, together with replication in independent cohorts incorporating amyloid and tau biomarker confirmation, will be essential to determine whether anxiety identifies individuals at increased risk for AD progression, represents an early clinical manifestation of preclinical disease, or both. If confirmed, assessment of anxiety may complement emerging blood-based biomarkers for identifying older adults at elevated risk during the preclinical stages of AD and inform future prevention strategies.

## CONCLUSION

Greater late-life global anxiety, but not worry or rumination, was selectively associated with higher plasma BD-p-tau181, a biomarker of AD pathology, in older adults enriched for anxiety-related symptoms. The absence of comparable associations with the non-BD form suggests that this relationship may reflect CNS processes rather than non-specific peripheral tau biology. Although the cross-sectional design precludes conclusions regarding causality or temporal sequence, these findings provide biological support for epidemiologic evidence linking anxiety to increased AD risk and underscore the importance of considering anxiety phenotypes as distinct rather than interchangeable constructs. Longitudinal follow-up incorporating cognitive outcomes in the RAW Brain study and amyloid/tau biomarker confirmation will be critical to determine whether elevated anxiety identifies individuals at increased risk for subsequent AD progression, represents an early neuropsychiatric manifestation of emerging pathology, or both. If confirmed, assessment of anxiety may complement blood-based biomarkers in identifying older adults at risk during the preclinical stages of AD and inform preventive intervention strategies.

## Authorship Statement

Conceptualization: APC, HTK, CA; Funding Acquisition: CA; Data Analysis: APC, DLT, HTK, CA; Writing original draft: APC, HTK, CA; Writing reviewing and editing: APC, HTK, MFF, NN, MMG, XZ, DLT, MAB, TKK, CA.

## Consent Statement

All participants gave written informed consent prior to starting study procedures. This study was approved by IRB at University of Pittsburgh.

## Previous Presentations

This work was presented at Pitt Psychiatry Annual Research Day 2026, Pittsburgh, PA, June 11, 2026.

### Data Sharing Statement

Data is available upon request from study principal investigator.

## Funding

This work was supported by National Institute of Mental Health grant (R01 MH108509 The RAW Brain – The Effect of Rumination, Anxiety and Worry on Aging and Dementia Risk). National Institute of Mental Health grant T32 MH019986.

## Declaration of Competing Interest

TKK has consulted for/served on advisory boards for Quanterix Corporation, SpearBio Inc., Neurogen Biomarking LLC, Alzheon, Grifols and Siemens Healthineers, has received honoraria from Cell Signaling Technology and Quebec Center for Drug Discovery, has been an investigator on studies supported by Fujirebio Inc. Genentech, Astellas Pharmaceuticals, Janssen Research Laboratories, SpearBio Inc., and Alamar Biosciences, has received royalties from Bioventix for the transfer of specific antibodies and assays to third party organizations, and is an inventor on patents and provisional patents regarding biofluid biomarker methods, targets and reagents/compositions, all outside the submitted work. The other authors report no competing interest.

## Data Availability

All data produced in the present study are available upon reasonable request to the authors

## Acknowledgements

We would like to thank participants for their participation in the study as well as study staff from the ARGO Neuroscience of Aging Research Group for their work and support.

## Notes

### Author Declarations

Ethics committee/IRB of University of Pittsburgh gave ethical approval for this work

