## Supplementary data for "Association between plasma brain-derived p-tau variants and anxiety phenotypes in older adults"

**Supplementary data**
**Table S1. Participant characteristics stratified by sex and symptom duration (<10 years vs. ≥10 years)**

**Table S1.** Participant characteristics stratified by sex and symptom duration (<10 years vs. ≥10 years)

| **Variable** | **High-RAW** N = 36[1] | **Low-RAW** N = 53[1] | **p-value**[2] | **Overall** N = 89[1] |
|---|---|---|---|---|
| ***Age, years, mean (SD)*** | 60.9 ± 8.1 | 66.6 ± 8.5 | **0.002** | 64.3 ± 8.8 |
| ***Sex, n, %*** | | | 0.255 | |
| Female | 28 (78%) | 34 (64%) | | 62 (70%) |
| Male | 8 (22%) | 19 (36%) | | 27 (30%) |
| ***Race, n, %*** | | | >0.999 | |
| Non-White | 6 (17%) | 8 (15%) | | 14 (16%) |
| White | 30 (83%) | 45 (85%) | | 75 (84%) |
| ***Ethnicity, n, %*** | | | 0.845 | |
| Hispanic or Latino | 1 (2.8%) | 0 (0%) | | 1 (1.1%) |
| Not Hispanic or Latino | 35 (97%) | 53 (100%) | | 88 (99%) |
| ***Education, years, mean (SD)*** | 16.3 ± 2.1 | 16.8 ± 2.2 | 0.330 | 16.6 ± 2.2 |
| ***APOE4 carrier status, n, %*** | | | 0.532 | |
| Carrier | 6 (17%) | 13 (25%) | | 19 (21%) |
| Non-carrier | 30 (83%) | 40 (75%) | | 70 (79%) |
| ***PSWQ (Worry), mean (SD)*** | 55.1 ± 10.7 | 31.5 ± 9.3 | **<0.001** | 41.1 ± 15.2 |
| ***RSQ (Rumination), mean (SD)*** | 47.4 ± 12.1 | 29.3 ± 5.7 | **<0.001** | 36.7 ± 12.5 |
| ***HARS (Anxiety), mean (SD)*** | 15.1 ± 7.0 | 5.4 ± 3.6 | **<0.001** | 9.3 ± 7.1 |
| ***Symptom duration, n, %*** | | | 0.444 | |
| Less than 10 years | 12 (33%) | 4 (57%) | | 16 (37%) |
| ≥10 years | 24 (67%) | 3 (43%) | | 27 (63%) |

[1] Mean ± SD; n (%)
[2] Wilcoxon rank sum test; Pearson's Chi-squared test
*Mean ± SD for continuous variables. n (%) for categorical variables.*
*p-values: Wilcoxon rank-sum test (continuous). Pearson Chi-squared test (categorical). With small or sparse cells consider Fisher's exact.*
*Symptom duration = max duration code across PSWQ, RSQ, HARS (codes 1–3 = <10 years; code 4 = ≥10 years).*

**Figure S1. Plasma tau measures do not differ between Low- and High-RAW groups.** Box plots compare normalized plasma levels of four plasma tau forms between participants stratified into Low-RAW and High-RAW groups (n = 89): (A) total-tau, (B) p-tau181, (C) p-tau217, and (D) p-tau231; unadjusted and adjusting (padj) for age, sex, and APOE4 carrier status.

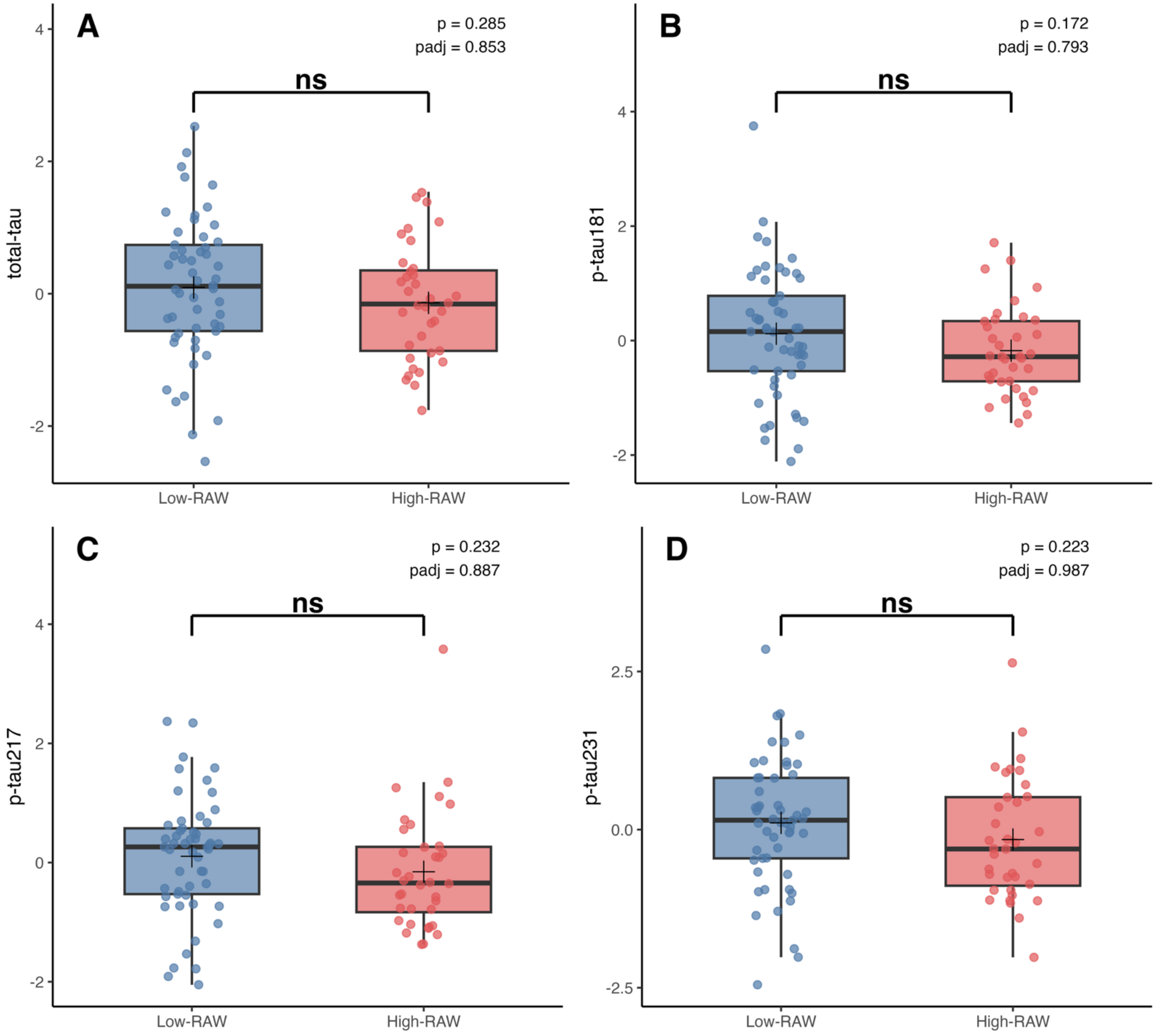

**Figure S2. Correlations between brain-derived and plasma phosphorylated tau versions.** (A) Pearson correlation coefficients (r) on standardized values between brain-derived and plasma phosphorylated tau versions. (B) Scatterplots showing the associations between brain-derived (BD) phosphorylated tau isoforms and their corresponding plasma measures. Each point represents an individual participant; the solid line represents the fitted linear regression line, with the shaded area indicating the 95% confidence interval.

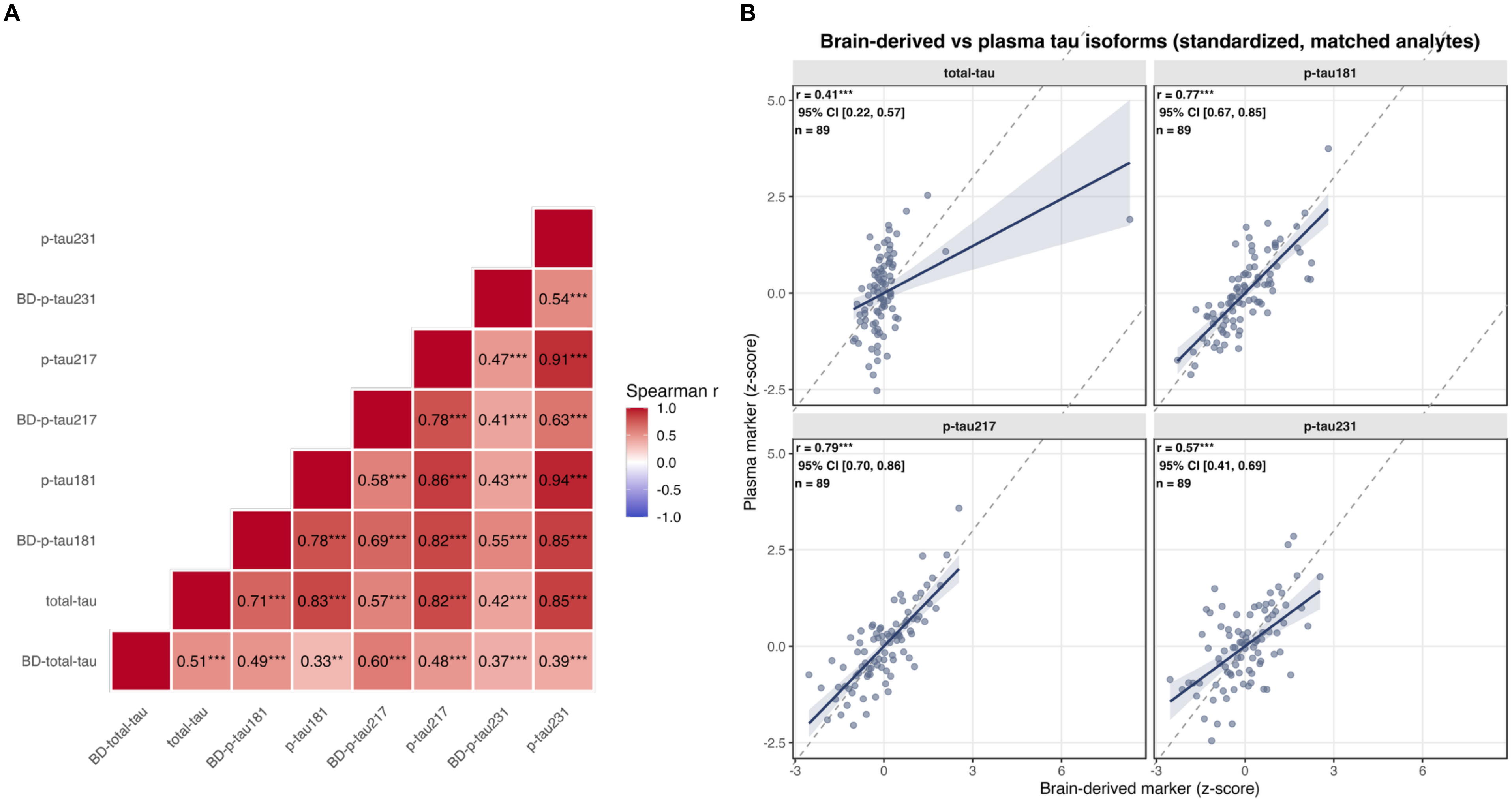

**Figure S3. Anxiety-associated phenotypes and their brain-derived tau correlates.** Each panel (A-C) corresponds to one brain-derived (BD) tau form (BD-tau, BD-p-tau217, and BD-p-tau231) associated with one or more anxiety phenotypes (worry, rumination, and global anxiety). Scatterplots of the BD-tau markers against anxiety phenotypes, with linear regression fit (solid line) and 95% confidence band (shaded). All models were adjusted for age, sex, and APOE4 status (N=89); standardized $\beta$, 95% CI, and FDR-adjusted p-value are shown.

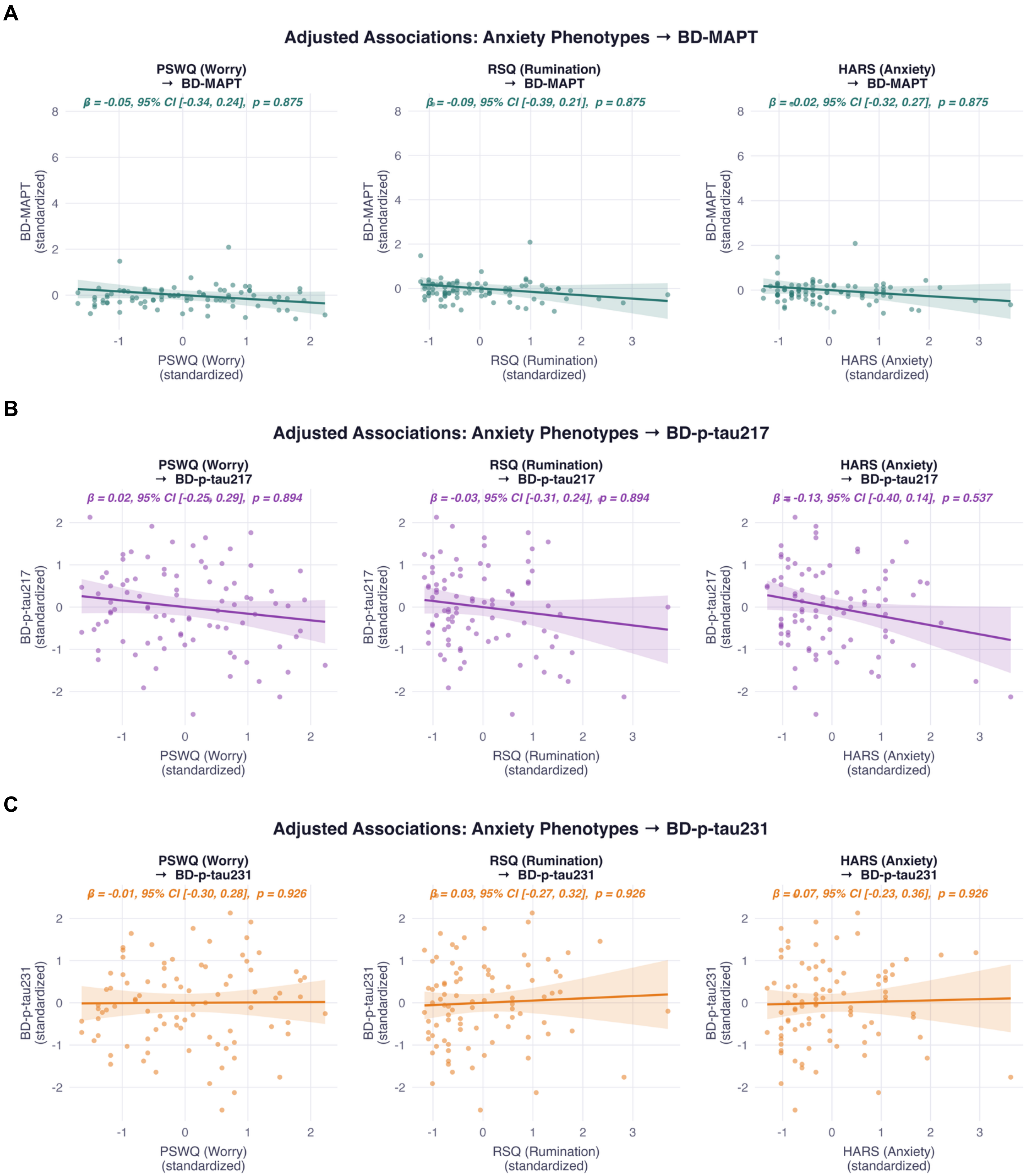

**Figure S4. Anxiety-associated phenotypes and their non-brain-derived tau correlates.** Each panel (A-D) corresponds to one non-brain-derived (non-BD) tau form (total-tau, p-tau181, p-tau217, and p-tau231) associated with one or more anxiety phenotypes (worry, rumination, and global anxiety). Scatterplots of the BD-tau markers against anxiety phenotypes, with linear regression fit (solid line) and 95% confidence band (shaded). All models were adjusted for age, sex, and APOE4 status (N=89); standardized β, 95% CI, and FDR-adjusted p-value are shown.

**A**

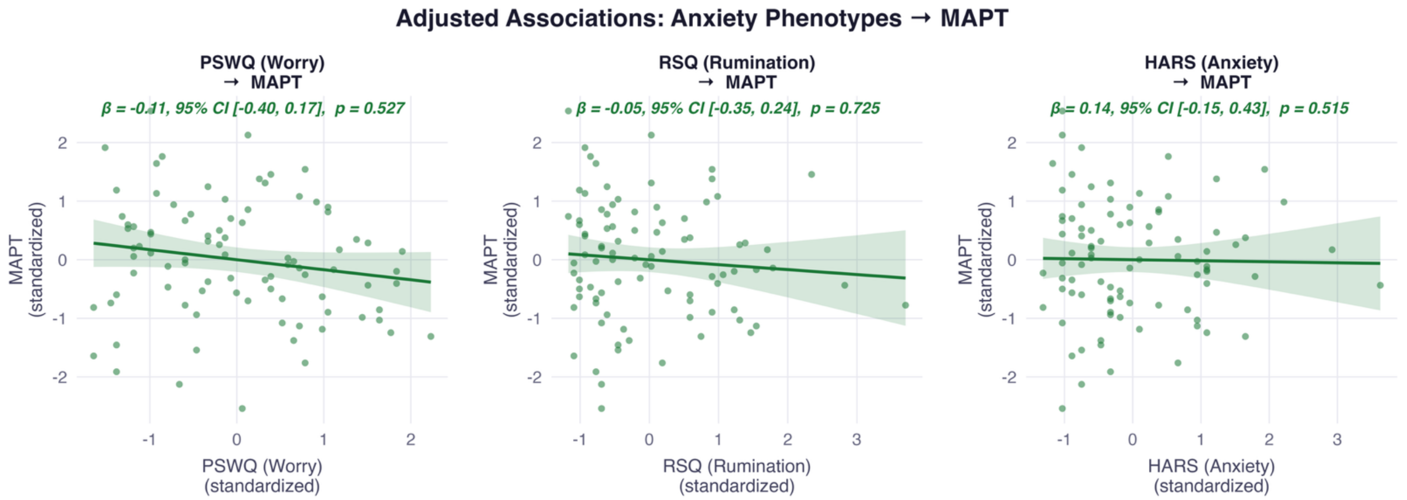


**B**

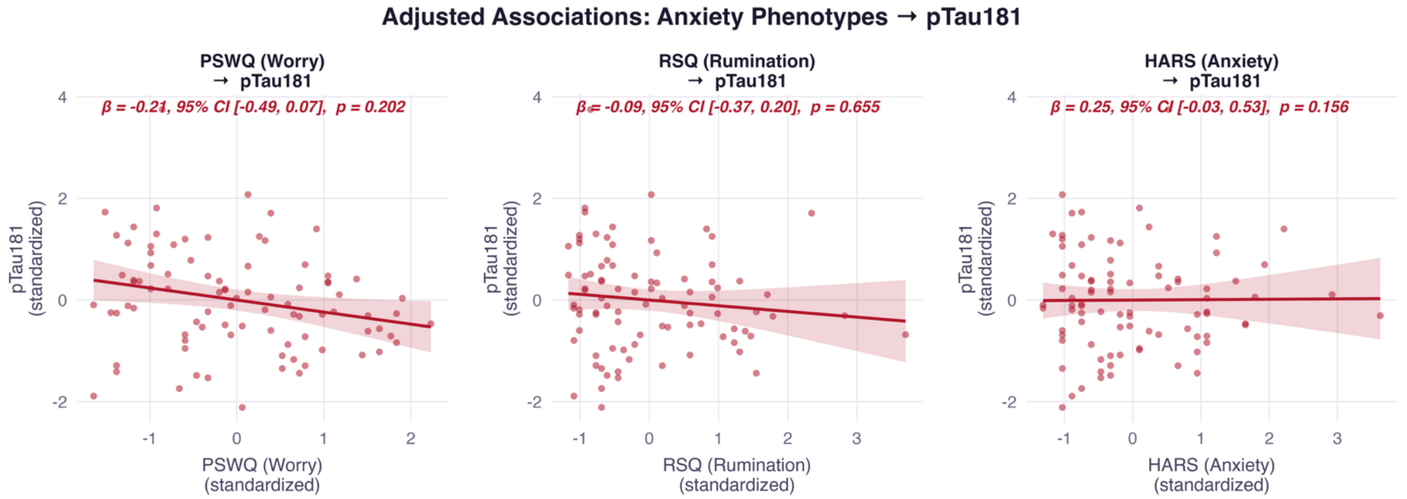


**C**

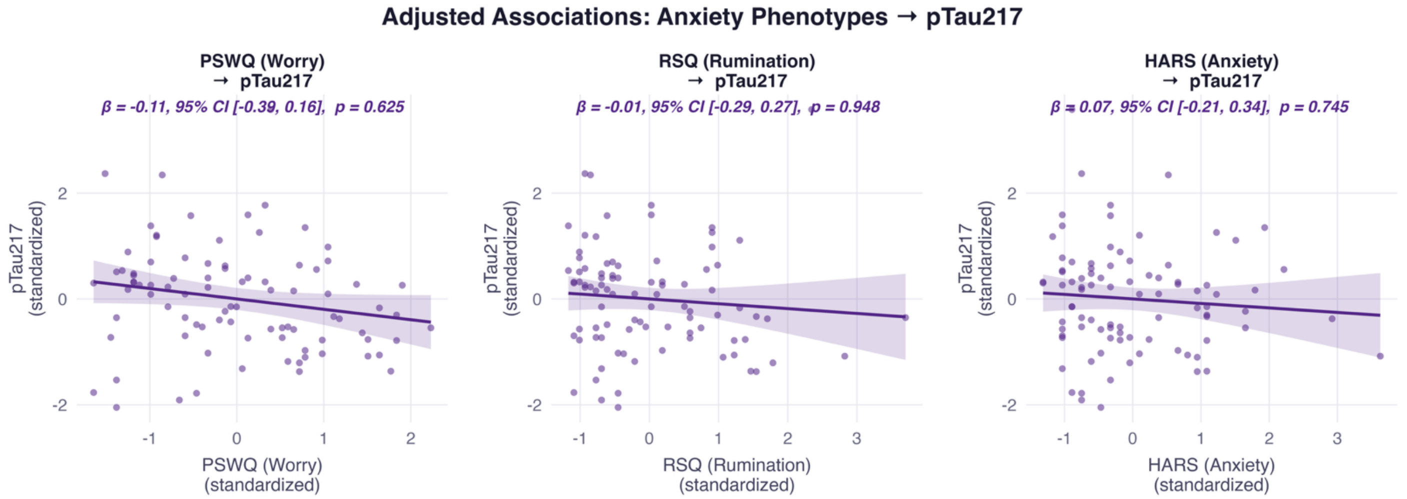

**D**

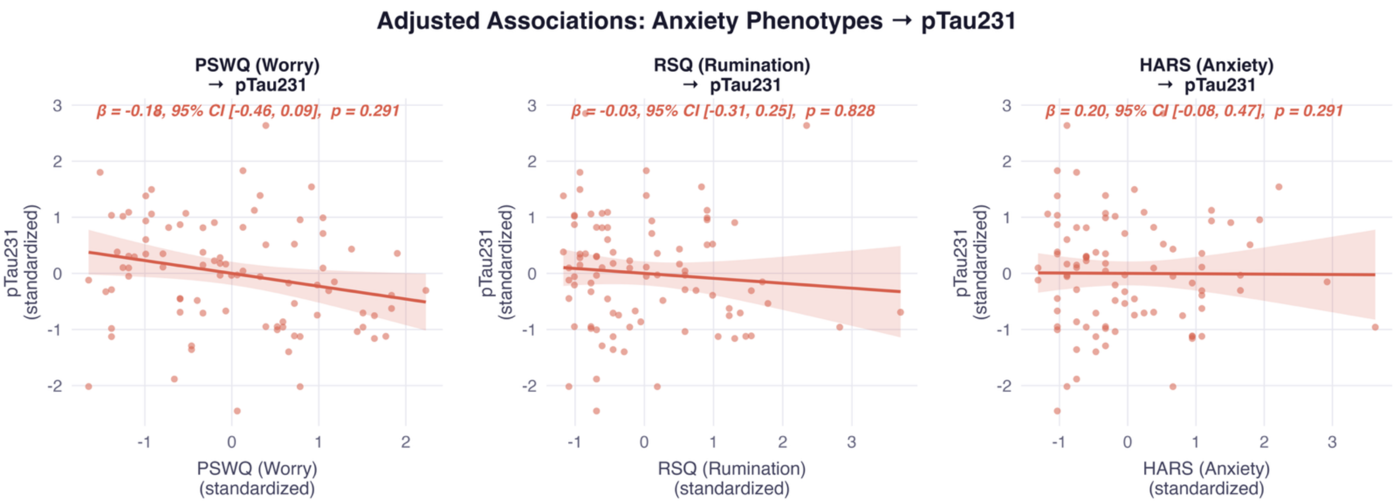
Adjusted Associations: Anxiety Phenotypes → pTau231
PSWQ (Worry) → pTau231
β = -0.18, 95% CI [-0.46, 0.09], p = 0.291
pTau231 (standardized)
PSWQ (Worry) (standardized)
RSQ (Rumination) → pTau231
β = -0.03, 95% CI [-0.31, 0.25], p = 0.828
RSQ (Rumination) (standardized)
HARS (Anxiety) → pTau231
β = 0.20, 95% CI [-0.08, 0.47], p = 0.291
HARS (Anxiety) (standardized)

**Figure S5. Stratified by sex: BD-tau and non-BD-tau forms**. Forest plots of standardized regression coefficients (β ± 95% CI) for the association between each anxiety phenotype (PSWQ, RSQ, HARS) and tau markers, stratified by sex (female, n = 62; male, n = 27). Models are adjusted for age and APOE4 carrier status; sex is not included as a covariate since it defines the stratification. Points to the right of the dashed reference line (β= 0) indicate a positive association between the anxiety phenotype and the tau marker within that sex. (A) Brain-derived (BD) tau markers: BD-MAPT, BD-p-tau181, BD-p-tau217, BD-p-tau231. (B) Non-BD (plasma) tau markers: MAPT, p-tau181, p-tau217, p-tau231. All associations shown here are non-significant after this FDR correction (ns).

**A**

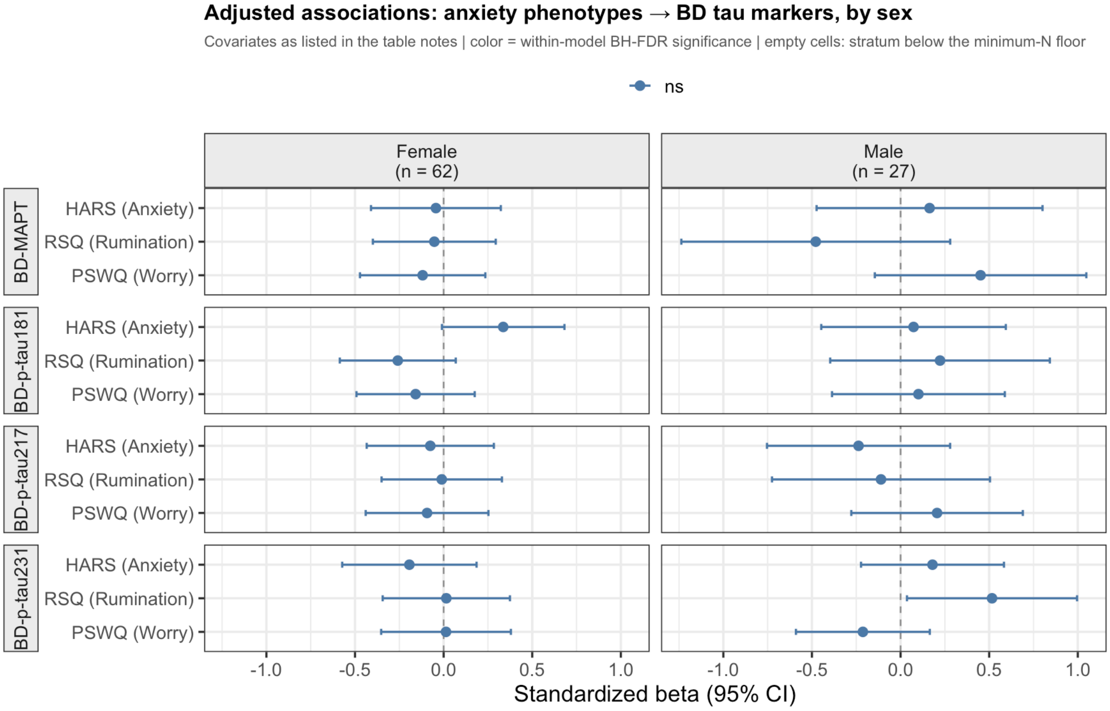


**B**

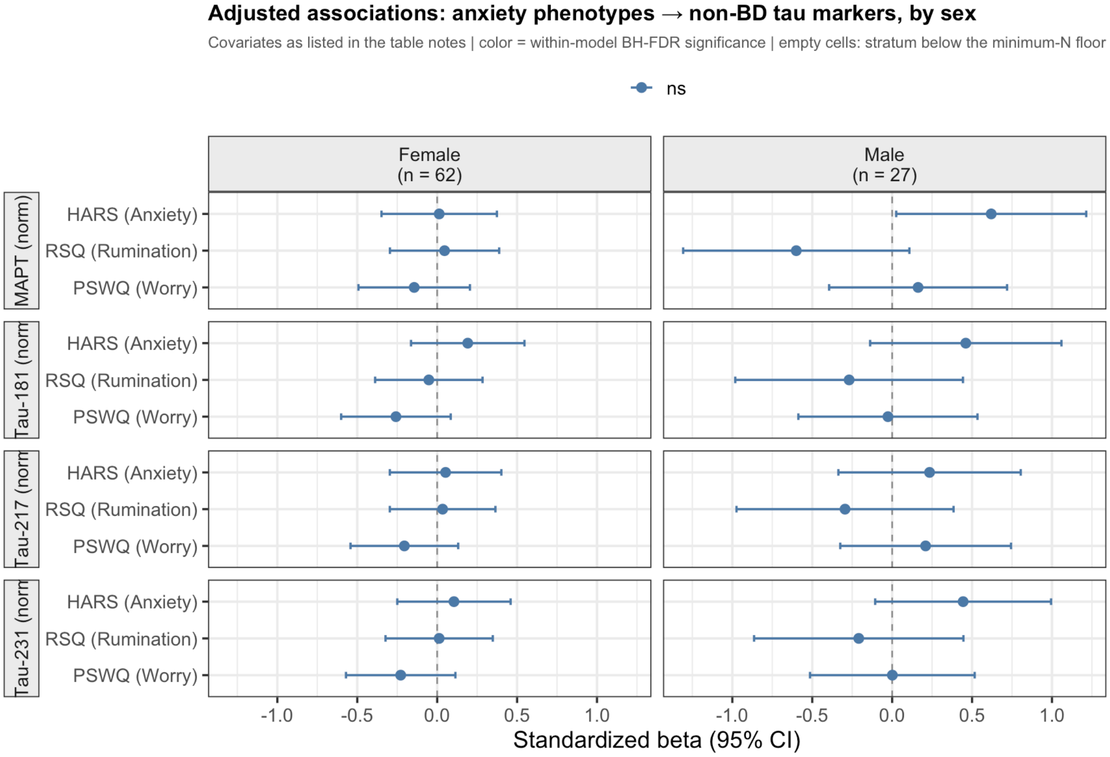

**Figure S6. Stratified by symptom duration: BD-tau and non-BD-tau versions.** Forest plots of standardized regression coefficients (β ± 95% CI) for the association between each anxiety phenotype (PSWQ, RSQ, HARS) and tau markers, stratified by symptom duration (<10 years, n = 16; ≥10 years, n = 27). Models are adjusted for age, sex, and APOE4 carrier status; symptom duration is not included as a covariate since it defines the stratification. Points to the right of the dashed reference line (β= 0) indicate a positive association between the anxiety phenotype and the tau marker within that duration stratum. (A) Brain-derived (BD) tau markers: BD-tau, BD-p-tau181, BD-p-tau217, BD-p-tau231. (B) Non-BD (plasma) tau markers: total-tau, p-tau181, p-tau217, p-tau231. All associations shown here are non-significant after this correction (ns): every confidence interval crosses zero.

**A**

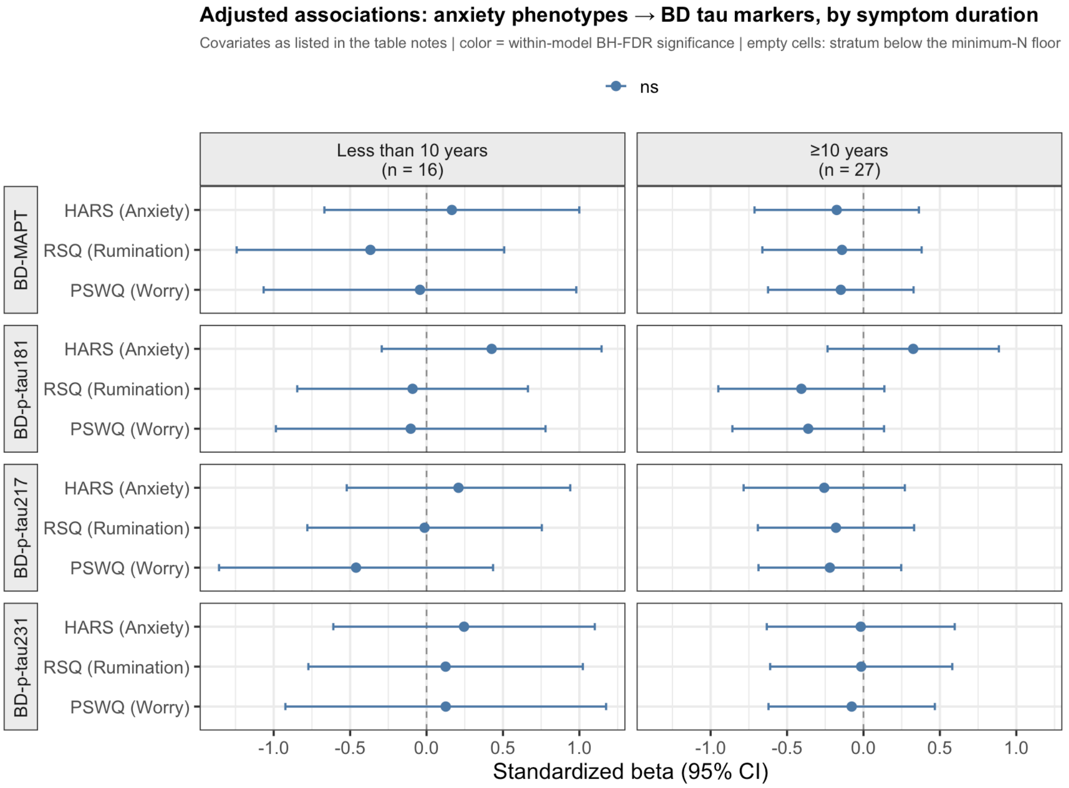


**B**

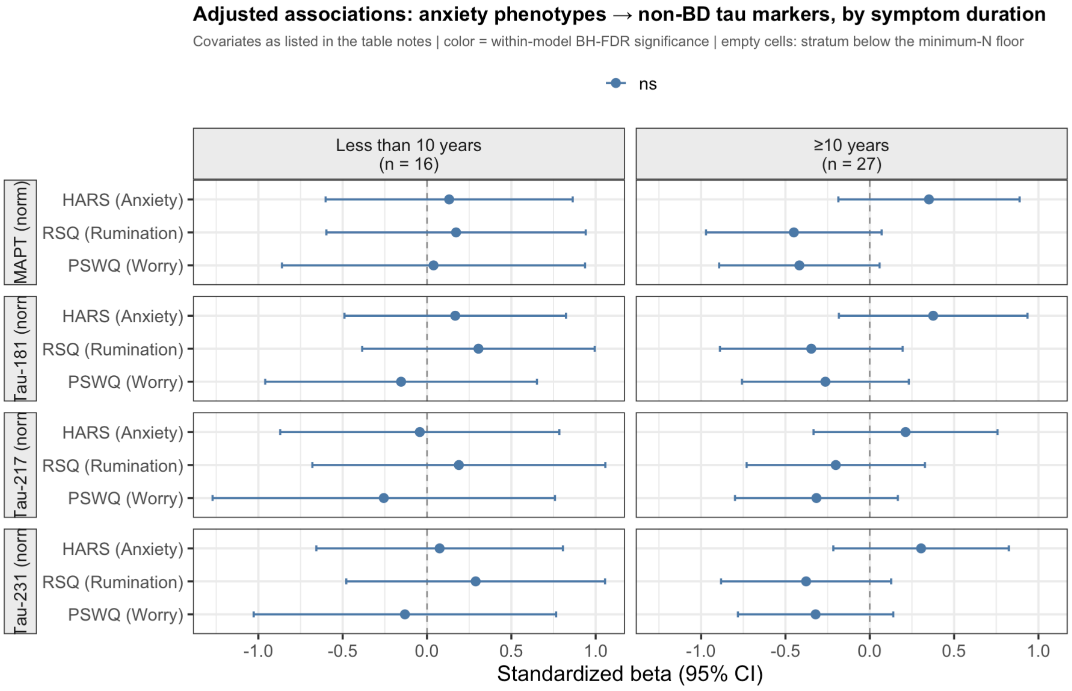
